# Shared Genetic Architecture of Psychosis, Mood, and Cognition in East Asian Ancestry

**DOI:** 10.64898/2026.06.01.26354666

**Authors:** Keane Lim, Tim van der Es, Jace Song, David M Howard, Jianjun Liu, Jimmy Lee, Chia-Yen Chen, Max Lam

**Author notes:** Merck & Co., Inc., Cambridge, MA 02141, USA. **Corresponding Author:** Max Lam, PhD, Institute of Mental Health, Singapore, 10 Buangkok View, Singapore 539747.

## Abstract

Genomic insights into psychiatric disorders remain heavily skewed toward European populations. In European-ancestry studies, educational attainment is typically negatively genetically correlated with major depression but paradoxically positively correlated with schizophrenia, raising the question of whether these relationships generalize across ancestries. We investigated whether this cross-trait architecture extends to East Asian ancestry (EAS). Using EAS GWAS summary statistics for major depressive disorder (MDD), schizophrenia (SZ), and educational attainment (EDU), we applied multi-trait (MTAG) and pleiotropy-informed (PLEIO) analyses to characterize shared genetic architecture across these traits. Across MTAG and PLEIO analyses, we identified 32 unique genome-wide significant loci (*p* < 5 × 10^−8^), including seven novel loci revealed in depression analysis that overlapped schizophrenia-associated signals – consistent with shared cross-trait architecture. Results reinforce a convergent risk architecture for affective and psychotic disorders in this population. Fine-mapping analyses prioritized variants mapping to candidate genes, including serine/threonine kinase *VRK2*, nominating targets for future follow-up. Cross-trait analyses supported a positive genetic relationship between EDU and MDD (*r*_*g*_ = 0.308, *p* = 9.63 × 10^−17^) in East Asian data, contrasting to the negative correlation typically observed in European ancestry. These findings suggest that the genetic relationship between educational attainment and psychiatric risk may not be fully transferable across ancestries. In an independent cohort of individuals at ultra-high risk for psychosis, MTAG-derived polygenic risk scores improved case-control discrimination relative to single-trait GWAS-based scores. These results underscore the importance of ancestry-specific genomic frameworks for interpreting cross-trait psychiatric architecture and improving polygenic prediction.

## Introduction

Major depressive disorder (MDD) and schizophrenia (SZ) are complex psychiatric conditions with lifetime prevalence rates of approximately 15% and 1%, respectively^1,2^, and high twin-based heritability estimates of about 30-50% for MDD and up to 80% for SZ^3,4^. These conditions often manifest with profound cognitive impairments^5,6^ and negative symptoms^7,8^ such as executive dysfunction and diminished motivation, respectively, which contribute to the chronic disability. Educational attainment (EDU), a heritable trait (20-40%), has emerged as a proxy phenotype for cognitive function, given its genetic correlation with cognition (*r*_g_ = ~0.6-0.85)^9,10^. Recent research has shown genetic correlations between EDU and both MDD and SZ, suggesting loci that influence educational success may also impact cognitive function in psychiatric disorders. The existing findings suggest that educational attainment could be a relevant trait for investigating shared psychiatric-cognitive genetic architecture, but it would remain an imperfect proxy for cognitive function in different contexts.

The genetic relationship between psychiatric disorders and cognitive traits shows a degree of complexity^11^. Prior reports suggest that the genetic architecture of EDU captures not only cognitive ability but also distinct biological or environmental pathways that may increase susceptibility to psychosis^12^. The relationship between EDU and MDD in European (EUR) cohorts has largely been characterized as negative (protective), where higher educational attainment correlates with lower depression risk^13,14^. Paradoxically, a positive genetic correlation was reported between EDU and SZ in EUR ancestries. Whether this generalizes across ancestries remains unclear, particularly in East-Asian (EAS) ancestry populations, where psychiatric GWAS are generally less powered, and the environmental context differs.

Genetic pleiotropy provides a framework for examining cross-trait relationships and potentially new insights from working with a multi-trait framework^15,16^. EDU captures a broad spectrum of cognitive abilities across the life course, offering a reliable proxy less influenced by transient clinical states^17,18^. Leveraging shared genetic architecture across correlated traits offer a way to improve discovery^19–21^, while testing whether cross-trait structure differs across ancestries, thereby furthering our understanding of the shared biological mechanisms underlying affective, psychotic, and cognitive domains.

Here, we aim to characterize the pleiotropic landscape of MDD, SZ, and EDU in East-Asian ancestry. Using EAS GWAS summary statistics for MDD, SZ and EDU, we will identify shared loci, compare cross-trait genetic relationships, and evaluate whether multi-trait models improve polygenic prediction in an independent EAS clinical-risk cohort. This study provides a comprehensive map of the shared biological mechanisms linking cognitive surrogates and psychiatric risk in EAS, laying a foundation for future studies exploring the underlying biological pathways. The findings hold potential for clinical advancements aimed at addressing both cognitive and psychiatric symptoms through more targeted interventions.

## Methods

A flowchart of the analyses described below is presented in Supplementary Fig. 1. Additional methods description is presented in Supplementary Information.

### Summary statistics quality control

The present study utilizes the latest East Asian ancestry GWAS summary statistics for educational attainment (N = 176,400; Chen et al)^21^, depression (N = 194,548; Giannakopouou et al)^20^, and schizophrenia (N = 58,140; Lam et al)^22^ (Supplementary Table 1). Quality control (QC) procedures were performed with the MungeSumstats tool v1.9.17^23^ to ensure consistency of summary statistics. The default QC parameters for MungeSumstats were applied. These include removing strand-ambiguous SNPs and non-biallelic SNPs, ensuring that all variants have non-zero effect sizes and standard errors, and standardizing the reference genome build (GRCh37 and dbSNP v.144), SNP ID, and the directionality of the effect allele. For downstream analysis, the Z-score and effective sample size were also estimated with MungeSumstats. QC procedures resulted in 4,229,706 SNPs shared across all three phenotypes (Supplementary Table 1). Genetic covariance intercept ranged between 0.003 to 0.046, indicating low sample overlap between traits (Supplementary Table 1). Input summary statistics and MTAG heritability metrics are also reported in Supplementary Table 1.

### Genome-wide analyses

Genome-wide analysis using MDD, SZ, and EDU GWAS was carried out with Multi-Trait Analysis of GWAS (MTAG v1.0.8)^24^, and Pleiotropic Locus Exploration and Interpretation using Optimal test (PLEIO v2.0)^25^ as a confirmatory approach. The MTAG framework enhances variant discovery by leveraging the genetic correlations among multiple traits. By incorporating correlation matrices for each trait, MTAG improves the accuracy and power of effect estimates, enabling the identification of genetic variants that may not be detectable through single-trait analyses. This approach enables a more comprehensive understanding of the shared genetic architecture across related traits, ultimately yielding more robust and insightful discoveries. The PLEIO framework employs a random-effects model that integrates genetic correlations and heritability estimates across multiple traits. This approach enhances the detection of pleiotropic variants—genetic variants that influence multiple phenotypes—by accounting for the shared genetic architecture among the traits. By leveraging this multi-trait information, PLEIO boosts the statistical power and precision of variant discovery, providing deeper insights into the biological mechanisms underlying complex traits.

### Loci identification

Independent genome-wide significant loci for each MTAG and PLEIO output were identified using the Functional Mapping and Annotation (FUMA, v1.6.1)^26^ pipeline. Default FUMA parameters of p-value < 5×10^−8^, LD r^2^ < 0.6, and East Asian 1000 genomes reference panel^27^ were set to identify independent SNPs and a second clumping of r^2^ < 0.1 to identify lead SNPs. To identify unique loci shared by MTAG and PLEIO outputs, we used a standardized approach to define their boundaries. Initially, loci were identified using FUMA, which employs clumping approaches to determine loci boundaries. However, due to subtle differences in association results and linkage disequilibrium (LD) patterns, the boundaries identified by FUMA may vary between MTAG and PLEIO analyses. To harmonize these loci’s boundaries, we standardized the loci by taking the overlapping upper and lower base-pair coordinates across the MTAG and PLEIO outputs. This means that the start and stop base coordinates for each locus identified were defined by the most inclusive boundaries from both analyses. By aligning these coordinates, we ensured that each locus is consistently defined, regardless of the method used to identify them. This harmonization process enables a more accurate and reliable comparison of loci shared between the MTAG and PLEIO outputs, facilitating the identification of pleiotropic variants with greater precision.

### Gene-based and gene-set analyses

The MAGMA v1.08^28^ gene-based and gene-set analyses, implemented within FUMA^26^, were conducted with 16,994 gene sets, including curated gene sets and GO terms from MsigDB v2023.1Hs^29^. Competitive gene set analysis was conducted, and the false discovery rate threshold (FDR < 0.05 Benjamin-Hochberg procedure) was applied to all tested gene sets. Both the gene-based and gene-set analyses were further filtered to present only results with three or more SNPs or genes, respectively. The g:Profiler^30^ was applied to four distinct gene sets derived from the MTAG and PLEIO analyses: PLEIO-only genes, which were genes significantly associated with the pleiotropic trait derived solely from the PLEIO analysis; MTAG-SZ-only genes, which were genes specifically associated with schizophrenia (SZ) in the MTAG analysis, but not significantly shared with major depressive disorder (MDD) or the overall pleiotropic set; MTAG-MDD-only genes, which were genes specifically associated with major depressive disorder (MDD) in the MTAG analysis, but not significantly shared with SZ or the overall pleiotropic set; and shared gene sets, which were genes found to be significantly associated across all three primary traits (pleiotropy, MTAG-SZ, and MTAG-MDD). For inclusion in the g:Profiler model, genes were filtered based on a nominal significance threshold of p < 0.05 from the respective genetic association analyses. Both approaches are complementary; MAGMA assesses whether a *predefined* set of genes (e.g., genes expressed in a specific brain region or cell type) exhibits a stronger association with the traits compared to other genes across the entire genome. Conversely, g:Profiler tests for the over-representation of genes within the identified sets across curated biological annotation resources, such as Gene Ontology (GO)^31^ terms (encompassing biological process, molecular function, and cellular component) and KEGG pathways^32^. This dual approach ensures a comprehensive interpretation, moving from genome-wide association to specific biological pathways and processes.

### Genetic correlation and heritability analyses

Global genetic correlation analyses were performed using Linkage Disequilibrium Score Regression (LDSC v1.0.1)^33,34^ on MDD, SZ, and EDU GWAS summary statistics and MTAG GWAS summary statistics to explore the relationships among the three phenotypes. The East Asian LD scores were based on the 1000 Genomes Phase 3 reference panel. SNP-based heritability was also estimated with LDSC.

### Fine-mapping analyses

Fine-mapping based on a single population sample was conducted with SuSiEx software implementation^35,36^ for the significant loci identified in the genome-wide analyses. The East Asian 1000 Genomes reference panel was used in fine-mapping. Next, putative causal variants from fine-mapping were annotated using Variant Effect Predictor (GRCh37, release 112, May 2024)^37^, and any previous significant genome-wide associations with other psychiatric or cognitive traits were identified using the GWAS catalog^38^.

### Polygenic risk score analyses

The polygenic predictive ability derived from the current genome-wide analyses for MTAG and PLEIO output was tested in a cohort of East Asian individuals at ultra-high risk for psychosis (n = 77) compared to healthy controls (n = 74). Details of this cohort (i.e., The Longitudinal Youth at Risk Study; LYRIKS) have been reported previously (N = 151)^39^.

Polygenic risk scores (PRS) were constructed using p-value thresholding and the Bayesian framework. The p-value thresholding method was implemented using PRSice2 v2.3.5^40,41^ to compute a weighted sum of risk alleles. The following parameters were applied: p-value clumping of r^2^ > 0.1 with 500 kb windows; East Asian individuals from the 1000 Genomes Phase 3 were utilized for linkage disequilibrium (LD) clumping of the variants; SNPs in the major histocompatibility complex (MHC; chr6:25–35 Mb) were removed due to the complex linkage disequilibrium pattern in this region. PRS was estimated for each participant for ten p-value thresholds (P_T_ ≤ 5 × 10^−08^, 1 × 10^−05^, 1 × 10^−04^, 1 × 10^−03^, 0.01, 0.05, 0.1, 0.02, 0.05, 1). The Bayesian framework was conducted with PRS-CS^42^. The East Asian 1000 Genomes LD reference panel and the global shrinking parameter were set to ⍰ = 1 × 10^-2 for all computations.

The PRS were standardized using means and standard deviations from the healthy controls. Logistic regression (Cases vs. Healthy controls) was performed using calculated PRS as predictors. In addition, the first 10 genetic principal components, age, and sex were included in each prediction model as covariates. A baseline comparison model was also carried out with PRS calculated from the original MDD, SCZ, and EDU GWAS summary statistics. The regression analyses were carried out in SPSS v23.

## Results

### Genome-wide analyses

Following quality control and harmonization, we integrated 4,229,706 common SNPs into the MTAG framework. This joint analysis substantially increased statistical power, identifying 37 genome-wide significant loci (P < 5 x 10-8) across the three traits (Supplementary Tables 1–2; Supplementary Figs. 2 and 4). We compared the MTAG GWAS results to the original MDD, SZ, and EDU GWAS (MDD vs MTAG-MDD; SZ vs MTAG-SZ and EDU vs MTAG-EDU), 7 loci identified by MTAG were novel to depression (MDD-MTAG) and 7 were novel to schizophrenia (SZ-MTAG). All 7 novel loci identified in MDD-MTAG overlapped with schizophrenia-associated loci from the original SZ GWAS (Fig. 1a; Supplementary Table 2). This complete overlap is consistent with substantial shared cross-trait architecture between affective and psychotic phenotypes in East Asian ancestry. Novel loci were defined as novel with respect to the input GWAS summary statistics (i.e., SCZ, MDD, and EDU). The list of reported loci from the input dataset that did not reach genome-wide significance in the current analysis is reported in Supplementary Table 3. Complementary analysis using PLEIO (based on 3,183,822 shared SNPs) further corroborated these findings, identifying 26 genome-wide significant loci (Supplementary Figs. 3–4). As above, PLEIO GWAS summary statistics were compared with the original SZ, MDD, and EDU GWAS summary statistics. Of these, 2 novel loci were unique to PLEIO output, while novel 7 loci overlapped with MDD-MTAG and novel 3 loci with SZ-MTAG. When combining results across the MTAG and PLEIO frameworks, a total of 32 unique novel genome-wide significant loci were identified across MTAG and PLEIO analyses (Table 1; Fig. 1b; Supplementary Table 2).

**Table 1.** Loci reaching genome-wide significance in MTAG or PLEIO analyses that were not genome-wide significant in the corresponding input East Asian GWAS. Note. SZ = Schizophrenia; MDD = Depression. Genes were annotated via Variant Effect Predictor

| Chr | Start | End | uniqID | rsID | BP | P | Dataset | Gene | Annotation |
| --- | --- | --- | --- | --- | --- | --- | --- | --- | --- |
| 1 | 97113731 | 97301707 | 1:97173906:C:T | rs12031518 | 97173906 | 4.75E-08 | SZ_MTAG | - | - |
| 1 | 179064875 | 179557130 | 1:179270314:C:T | rs12031894 | 179270314 | 4.06E-08 | SZ_MTAG | SOAT1 | Intron |
| 1 | 179064875 | 179557130 | 1:179270314:C:T | rs12031894 | 179270314 | 1.31E-08 | PLEIO | SOAT1 | Intron |
| 2 | 27852150 | 28293299 | 2:28024642:C:T | rs13019037 | 28024642 | 4.34E-08 | PLEIO | RBKS; MRPL33 | Intron; NMD transcript |
| 2 | 57947592 | 58506441 | 2:58380167:A:G | rs35844949 | 58380167 | 1.08E-13 | MDD_MTAG | VRK2 | Intron |
| 2 | 57951208 | 58843588 | 2:58380167:A:G | rs35844949 | 58380167 | 2.19E-21 | PLEIO | VRK2 | Intron |
| 2 | 188421099 | 188421099 | 2:188421099:C:T | rs145763517 | 188421099 | 1.39E-08 | PLEIO | TFPI; AC007319.1 | Upstream gene; Downstream gene |
| 2 | 200720420 | 201247497 | 2:201176071:C:T | rs17592552 | 201176071 | 9.71E-15 | PLEIO | SPATS2L | Intron |
| 2 | 201085868 | 201247497 | 2:201176071:C:T | rs17592552 | 201176071 | 2.75E-10 | MDD_MTAG | SPATS2L | Intron |
| 4 | 66459249 | 66542686 | 4:66504803:A:G | rs62297721 | 66504803 | 3.06E-08 | SZ_MTAG | EPHA5 | Intron |
| 5 | 45136050 | 46405055 | 5:45305615:A:G | rs6874127 | 45305615 | 3.10E-08 | SZ_MTAG | HCN1 | Intron |
| 5 | 140751128 | 141017339 | 5:140853856:C:T | rs11956411 | 140853856 | 2.42E-08 | SZ_MTAG | PCDHGA12 | Intron |
| 5 | 140761777 | 141017339 | 5:140853856:C:T | rs11956411 | 140853856 | 3.86E-08 | PLEIO | PCDHGA12 | Intron |
| 6 | 165063590 | 165178845 | 6:165120422:A:G | rs1812595 | 165120422 | 3.58E-11 | PLEIO | - | - |
| 6 | 165066062 | 165178845 | 6:165100721:C:T | rs6455956 | 165100721 | 1.35E-08 | MDD_MTAG | - | - |
| 7 | 137019445 | 137085250 | 7:137070207:C:T | rs983708 | 137070207 | 1.36E-12 | PLEIO | DGKI | 3 prime UTR; Downstream gene |
| 7 | 137039670 | 137085250 | 7:137070207:C:T | rs983708 | 137070207 | 4.95E-08 | MDD_MTAG | DGKI | 3 prime UTR; Downstream gene |
| 8 | 26133566 | 26279173 | 8:26242272:G:T | rs117325001 | 26242272 | 1.60E-08 | SZ_MTAG | BNIP3L; SDAD1P1 | Intron; Upstream gene |
| 10 | 64848159 | 64892322 | 10:64862477:C:T | rs112222723 | 64862477 | 3.56E-08 | SZ_MTAG | - | - |
| 10 | 64848159 | 65309777 | 10:64849772:A:C | rs111830767 | 64849772 | 1.19E-08 | PLEIO | - | - |
| 10 | 64875311 | 65309777 | 10:64878562:C:T | rs6479881 | 64878562 | 4.40E-08 | MDD_MTAG | - | - |
| 12 | 123223920 | 123909466 | 12:123644043:C:T | rs61041384 | 123644043 | 4.62E-14 | PLEIO | MPHOSPH9 | Intron |
| 12 | 123409694 | 123747783 | 12:123541606:C:T | rs10773921 | 123541606 | 1.94E-09 | MDD_MTAG | PITPNM2 | Intron |
| 18 | 77583797 | 77702976 | 18:77633629:A:G | rs56181785 | 77633629 | 3.93E-11 | PLEIO | KCNG2 | Intron |
| 18 | 77587927 | 77647406 | 18:77633629:A:G | rs56181785 | 77633629 | 7.54E-09 | MDD_MTAG | KCNG2 | Intron |

**Fig. 1.**
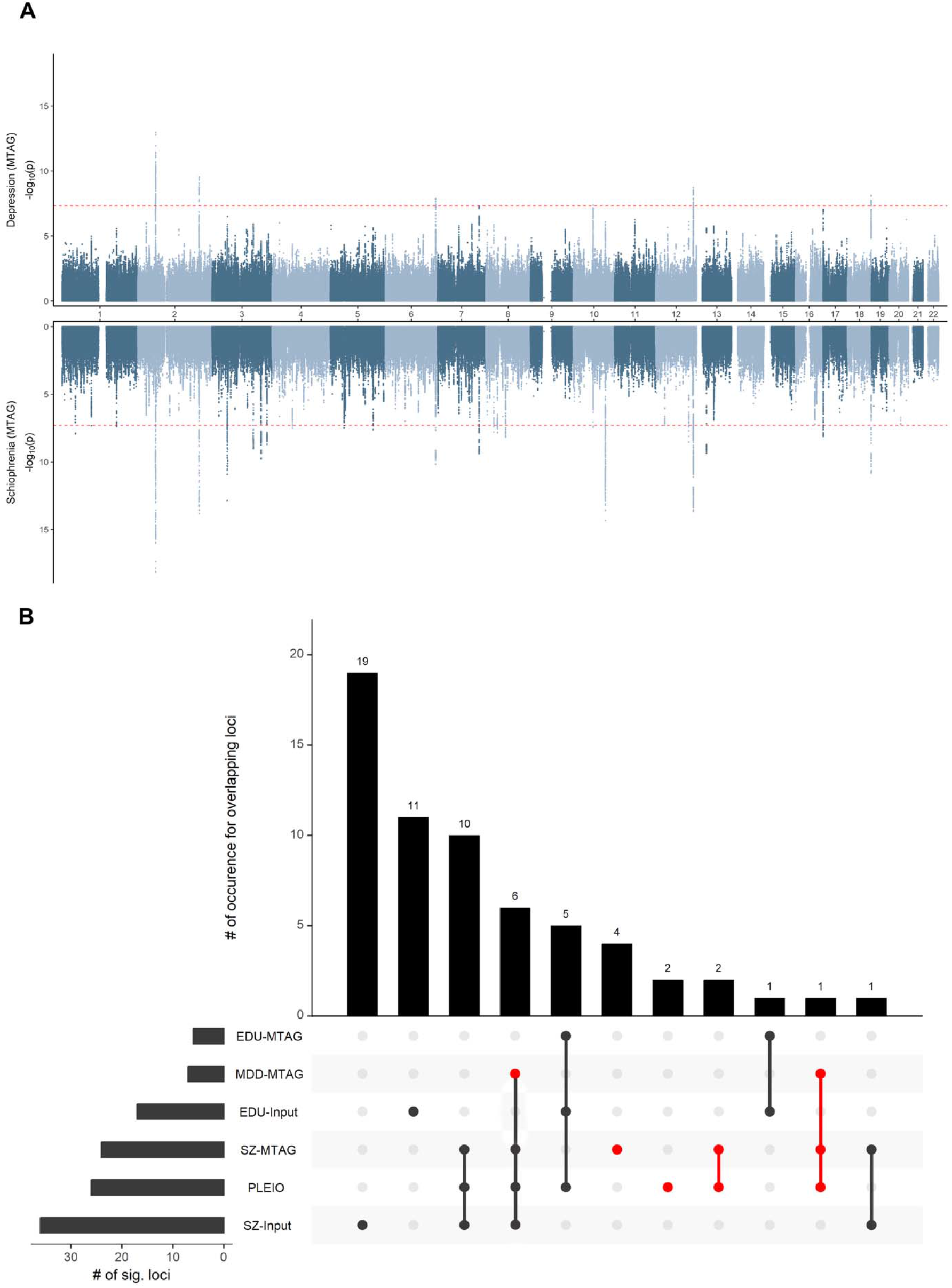
Genome-wide loci for MTAG depression and MTAG Schizophrenia. (A) Top panel: Manhattan plot of genome-wide association for depression via MTAG. Bottom panel: Manhattan plot of genome-wide association for schizophrenia via MTAG. The red dotted line represents genome-wide significance of p = 5×10^−8^. (B) Visualisation for overlapping genome-wide significant loci between each trait. The red dots represent novel genome-wide loci for the indicated trait. EDU-MTAG = Educational attainment MTAG output; MDD-MTAG = Depression MTAG output; SZ-MTAG = Schizophrenia MTAG output; EDU-Input = Discovery input dataset by Chen et al. (2024); SZ-Input = Discovery input dataset by Lam et al. (2019). Novel loci were defined relative to genome-wide significance in the corresponding input East Asian GWAS.

To dissect the specific contributions of each trait and assess the stability of these associations, we performed pairwise MTAG analyses (ie., MDD+SZ, MDD+EDU, EDU+SZ) in addition to the results presented above. Pair-wise analysis (Supplementary Table 2; Supplementary Fig. 5) revealed similar results, with the exception of 3 EDU loci, which showed attenuated significance in the three-way model, but remained significant in the pairwise analysis, albeit weaker, compared to the input EDU summary statistics. Conversely, the pairwise analysis captured two additional schizophrenia loci, further illustrating the utility of exploring varying trait combinations to maximize discovery.

### Gene-based and gene-set analyses

To characterize the biological function of the identified associations and to map the novel genome-wide significant SNPs to genes, we performed gene-based analysis using MAGMA. A total of 17,473 and 15,899 autosomal genes were mapped to the MTAG (MDD and SZ), and PLEIO output were prioritized, respectively (Supplementary Table 4-6). The MAGMA gene-set analyses revealed no significant pathways after FDR correction (Supplementary Table 7-9).

Functional enrichment analysis with g:Profiler shows that the shared core (PLEIO ⍰ SZ ⍰ MDD) converges on neurodevelopment, cell adhesion, and synaptic organization localized to cortical/hippocampal neuronal compartments, supporting a shared circuit-assembly and synaptic plasticity substrate across disorders (Supplementary Table 10). The PLEIO-only genes emphasize upstream transcriptional/chromatin regulation, cytoskeletal remodeling, circadian and neuroendocrine–metabolic pathways, with tau/ataxia signals implicating microtubule and proteostasis-related pleiotropic effects. The MTAG-SZ-only enrichment highlights Ca^2+^/ion transport, postsynaptic organization, phosphatase–HDAC regulation, and proteostasis, with dopaminergic synapse enrichment aligning with canonical SZ biology. The MTAG-MDD-only genes center on stress-responsive synaptic plasticity, postsynaptic receptor regulation, and RNA granule–mediated translational control, anchored by glutamatergic synapse pathways.

### Genetic correlation and heritability analyses

To place the MTAG results in context, we first examined genetic correlations in the input East Asian GWAS (Supplementary Table 11). We then compared these with the MTAG-derived outputs, which sharpened cross-trait relationships and increased discovery power. A robust positive genetic correlation was observed for EDU-MTAG with both psychiatric phenotypes, MDD-MTAG (*r*_*g*_ = 0.308, *p* = 9.63 × 10^−17^) and SZ-MTAG (*r*_*g*_ = 0.343, *p* = 5.08 × 10^−28^). A positive genetic correlation was also observed for MDD-MTAG and SZ-MTAG (*r*_*g*_ = 0.443, *p* = 6.17 × 10^−32^). This pattern indicates that in EAS ancestry, genetic variants associated with higher education are aligned with increased risk for both mood and psychotic disorders. Full genetic correlation results are reported in Supplementary Table 11. MTAG analysis yielded substantial SNP-based heritability estimates of *h*^2^ = 0.192 (*SE* = 0.012); *h*^2^ = 0.486 (*SE* = 0.027); and *h*^2^ = 0.00895 (*SE* = 0.0057) on the observed scale for MDD-MTAG, SZ-MTAG, and EDU-MTAG, respectively, confirming that the joint analysis effectively captured the polygenic signal (Supplementary Table 1).

### Fine-mapping analyses

To further refine the novel genomic loci identified, fine-mapping analyses for putative causal variants were performed with SuSiEx. We focused on identifying high-confidence causal variants, SNPs with genome-wide significance (p < 5 × 10^−8^), and a cumulative posterior inclusion probability of 90% credible sets were presented in Supplementary Tables 12-14 and Supplementary Figures 6-7. Rigorous filtering identified 5 credible sets for MDD-MTAG and 1 for SZ-MTAG. Function annotation of the top putative causal SNP in the MTAG results was carried out with Variant Effect Predictor (Supplementary Table 15-17), and a lookup in the GWAS catalog was performed to identify the mapped genes with previously reported GWAS trait associations (Supplementary Table 15). Functional annotation revealed that these credible sets map primarily to intronic and downstream regulatory regions (Supplementary Tables 15–17). Most notably, fine-mapping prioritized variants within *VRK2* and *FANCL*. While these variants are non-coding, their localization within *VRK2*—a gene encoding a serine/threonine kinase involved in neuronal proliferation—is particularly significant. Given prior evidence linking VRK2 to neurodevelopmental and psychiatric phenotypes, its prioritization highlights a plausible candidate for future functional investigation

### Polygenic risk score analyses

To evaluate whether multi-trait models improved polygenic discrimination in an independent East Asian clinical-risk cohort, we tested PRS performance in individuals at ultra-high risk for psychosis versus healthy controls (Supplementary Table 18). We compared these scores with baseline PRS derived from standard single-trait GWAS to quantify the added value of the MTAG framework. MTAG-derived scores yielded significant improvements in case-control discrimination compared to baseline PRS. The MTAG-SZ score achieved the highest predictive accuracy (Nagelkerke R^2^ = 0.138, P = 1.0 × 10^−4^), associated with a greater than two-fold increase in risk per standard deviation increase in score (OR = 2.23; 95% CI 1.49–3.34) (Fig. 2). Similarly, the MTAG-MDD score demonstrated robust prediction (R^2^ = 0.097, P = 0.001), with a significant Odds Ratio of 1.99 (95% CI 1.32–2.99). While the composite PLEIO score also achieved nominal significance (R^2^ = 0.069, P = 0.004, p_T_ = 0.5, OR = 1.69, 95% CI = 1.18-2.41), the trait-specific MTAG scores provided superior predictive power. (Supplementary Fig. 8). These results confirm that leveraging genetic correlations from large-scale datasets significantly enhances risk stratification in smaller, clinically severe cohorts. Because the external cohort was modest in sample sizes, these analyses should be interpreted as independent benchmarking rather than definitive clinical validation.

**Fig. 2.**
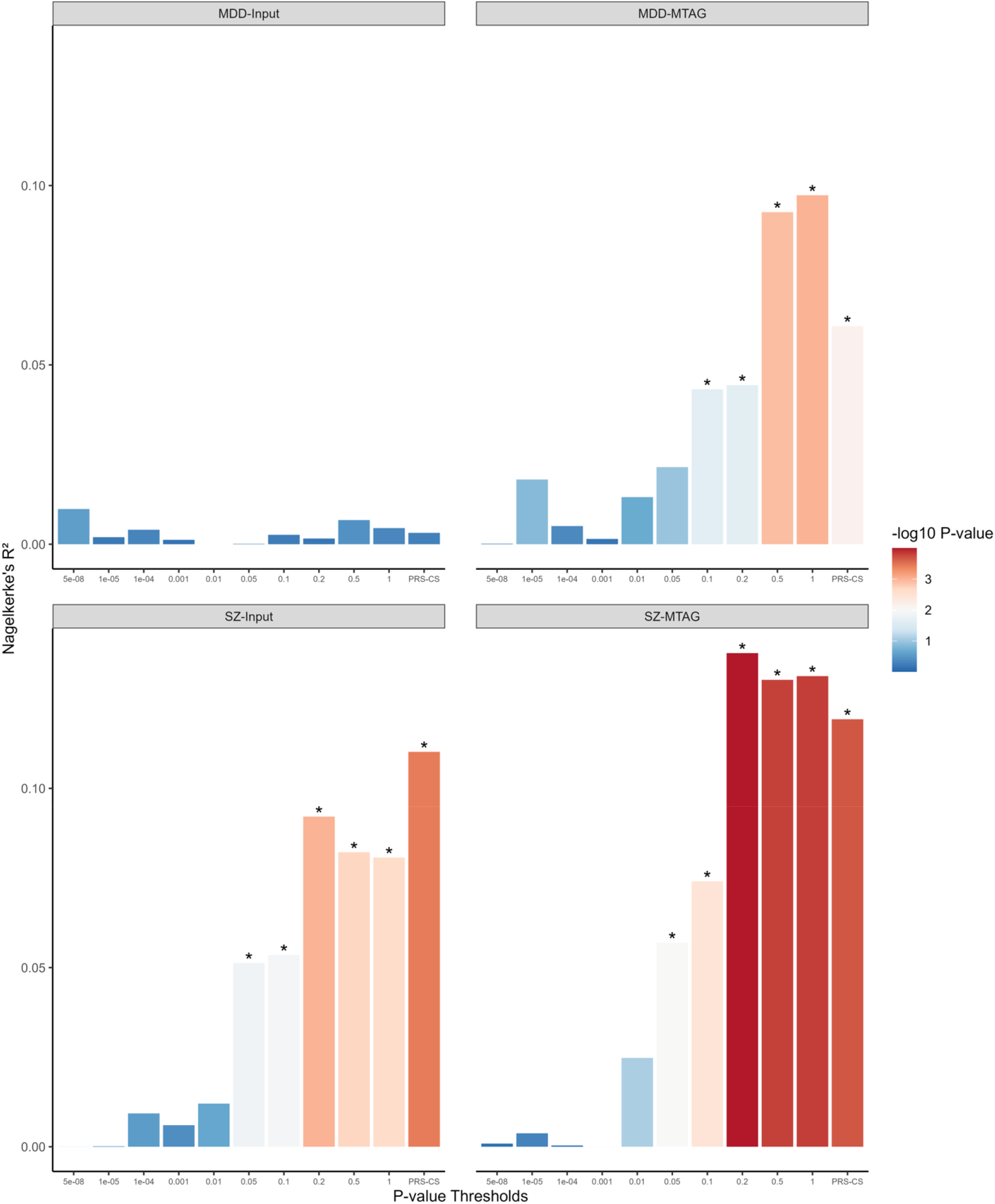
Polygenic prediction in individuals at ultra risk for psychosis versus healthy controls. *Note*. Polygenic score performance in an independent East Asian ultra-high-risk cohort (77 UHR cases, 74 controls). MTAG-derived scores are compared with matched single-trait GWAS-based scores; threshold-specific exploratory analyses are presented in Supplementary Figure 8. MDD-Input = The discovery input dataset by Giannakopoul et al. (2021), with quality control procedures applied via MungeSumstats; SZ-Input = The discovery input dataset by Lam et al. (2019), with quality control procedures applied via MungeSumstats. MDD-MTAG = Depression MTAG output; SZ-MTAG = Schizophrenia MTAG output. *p < 0.05.

## Discussion

### Pleiotropic discovery

This study extends cross-trait psychiatric genomic analysis to East Asian ancestry using large summary statistics for depression, schizophrenia, and educational attainment. By leveraging shared architecture, the multi-trait framework increased discovery relative to single-trait analyses and identified loci consistent with overlap between affective and psychotic phenotypes in this population. We report the results of genome-wide multi-trait analysis of MDD, SZ, and EDU, for East Asian ancestry individuals, to identify pleiotropic mechanisms underlying these traits. We identified 32 unique loci that were significantly associated with at least one trait. Crucially, we demonstrate a convergent genetic architecture between affective and psychotic disorders: all 7 novel depression loci displayed pleiotropic overlap with schizophrenia. This complete overlap validates the biological reality of a shared genetic substrate for these conditions in East Asian ancestry. While pairwise analysis revealed additional genetic signals, these loci were less genome-wide significant than the input trait and were not significant in the 3-way analysis. This suggests that by adding more traits in an MTAG model, pleiotropic loci that index cross-trait mechanisms may be more distinctly distinguished from noise that is often captured in lower-dimensional analyses.

### Pathway and functional analysis

Formal MAGMA gene-set analyses did not survive multiple-testing correction, so pathway interpretation should be considered exploratory. The g:Profiler enrichment converges on biological interpretations of prioritized gene-sets. The top pathways implicated in MDD included translational control pathways, which are crucial for the initiation of protein synthesis and cellular growth and response to environmental stress; and the gap junction assembly and function pathway, which are specialized cell-cell connections that allow direct communication between adjacent cells. The dysregulation in these pathways has been previously reported in autism spectrum disorder, schizophrenia, depression, and cognition^43,44^. These findings also corroborated with the genes indexing the top credible SNPs, previously implicated in genome-wide studies on schizophrenia, depression, and cognitive performance (Supplementary Table 15).

Fine-mapping and downstream annotation highlighted candidate genes with plausible relevance to shared affective–psychosis biology. *VRK2* is notable because it encodes a serine/threonine kinase previously prioritized as a potentially druggable schizophrenia-associated target through gene-prioritization analyses^45^, and because expression studies link altered *VRK2* expression to schizophrenia, bipolar disorder, and depression-related phenotypes^46,47^. Although structural and inhibitor studies support the broader pharmacological tractability of VRK-family kinases^48–52^, the present genetic analyses do not establish therapeutic directionality, target mechanism, or clinical actionability. Additional pharmacological annotation of *EPHA5* and *BNIP3L* points to neurodevelopmental Eph/ephrin signaling^53–60^ and mitochondrial stress/mitophagy biology^61–71^, respectively. These findings should therefore be interpreted as hypothesis-generating candidates for functional validation rather than evidence of ready therapeutic targets (see Supplementary Information).

### Genetic correlation

One of the main ancestry-specific observations in this study is the positive relationship between educational attainment and depression in East Asian data, contrasting with the more commonly reported negative relationship in European-ancestry cohorts^13^. The relationship between educational attainment and schizophrenia is more complex and has previously shown paradoxical positive correlations in European studies, making the depression finding the clearer divergence.

These mechanisms remain speculative and warrant direct study with better environmental and social phenotyping. First, the genetic architecture of MDD is highly polygenic, due to heterogeneity in MDD definitions across cohorts and ancestries. In turn, this may influence the directionality of genetic correlation at specific genomic segments, thereby affecting the overall global correlation. Second, non-genetic factors such as geographical regions may be expressed and modified differently through gene-environment interactions^20,72^. For instance, the ‘protective’ effect of education is not a universal biological constant but rather contingent on environmental and cultural contexts. We hypothesize that this divergence reflects distinct sociogenomic interactions. Differences in cultural norms^73–75^ or social factors, such as socioeconomic status, may also affect access to appropriate interventions and educational opportunities, thereby shaping the relationship between these two phenotypes. This finding challenges the generalizability of Eurocentric psychiatric models and underscores the need to interpret genetic correlations through an ancestry-aware lens. Further evaluation on the phenotype ascertainment bias, degree of polygenic overlap, and local genetic correlation, or the influence of cultural context are needed to dissect this differing phenomenon observed in East Asian and European ancestries.

### PRS implications

The PRS analysis suggests that multi-trait models may improve polygenic discrimination in clinically enriched East Asian samples relative to single-trait baselines. This result is encouraging, but the modest sample size and case-control design mean it should be viewed as external benchmarking rather than clinical implementation evidence. Further replication in larger prospective cohorts will be needed to determine whether these gains translate into robust risk stratification. By integrating correlated phenotypes, MTAG significantly improved the discrimination of individuals at Ultra-High Risk (UHR) for psychosis. Specifically, improved prediction was observed when MDD and SZ was applied, but not for EDU. This specificity implies that MTAG successfully captured the shared biological pathways driving general psychopathology (often conceptualized as the ‘p-factor’). Given that the UHR state is increasingly recognized as a broad syndrome of pluripotential risk rather than a specific precursor to psychosis alone^76,77^, our results validate this shift. The ability of our pleiotropy-informed scores to better classify these high-risk youth suggests that UHR status is fundamentally a manifestation of severe, shared genomic vulnerability across the affective-psychotic spectrum.

### Limitations

Several limitations should be noted. First, educational attainment is an imperfect proxy for cognition and captures social as well as cognitive factors. Second, MTAG improves power by borrowing information across correlated traits, hence the loci revealed in depression analyses may reflect shared cross-trait architecture rather than depression-specific biology. Third, formal pathway analyses did not survive multiple-testing correction, making downstream enrichment results exploratory. Fourth, the external PRS analysis was conducted in a modestly sized UHR cohort and should be interpreted as independent benchmarking rather than clinical validation. Finally, the candidate genes prioritized here, including *VRK2*, remain of nominal statistical significance, hence requiring further functional follow-up.

## Conclusion

In summary, cross-trait genomic analysis in East Asian ancestry identified shared loci across depression, schizophrenia, and educational attainment, and highlighted an ancestry-specific positive relationship between educational attainment and depression. Multi-trait modeling also improved polygenic discrimination in an independent East Asian clinical-risk cohort. These findings support the need for ancestry-specific psychiatric genomic frameworks and provide candidate loci and predictive models for future functional and prospective validation. Further work comparing cross-ancestry cross-trait analyses is needed to demonstrate that equitable precision psychiatry requires not just the application of existing tools to diverse populations, but also the fundamental characterization of population-specific genomic landscapes.

## Data Availability

All data produced in the present study are available upon reasonable request to the authors

## Funding acknowledgement

This study was supported by Singapore Ministry of Health’s National Medical Research under the Centre Grant Programme (Grant No.:NMRC/CG1/005/2021-IMH). We also acknowledge the National Research Foundation Singapore under the National Medical Research Council Translational and Clinical Research Flagship Programme (Grant No.: NMRC/TCR/003/2008) for the LYRIKS cohort. DMH is funded by an MRC Career Development Award (MR/Y011112/1).

## References

1. Insel, T. R. Assessing the economic costs of serious mental illness. Am. J. Psychiatry 165, 663–665 (2008).

2. Vos, T. et al. Global, regional, and national incidence, prevalence, and years lived with disability for 328 diseases and injuries for 195 countries, 1990–2016: a systematic analysis for the Global Burden of Disease Study 2016. Lancet 390, 1211–1259 (2017).

3. Hilker, R. et al. Heritability of schizophrenia and schizophrenia spectrum based on the nationwide Danish Twin Register. Biol. Psychiatry 83, 492–498 (2018).

4. Tabrizi, F. et al. Heritability and polygenic load for comorbid anxiety and depression. Transl. Psychiatry 15, 98 (2025).

5. Rock, P. L., Roiser, J. P., Riedel, W. J. & Blackwell, A. D. Cognitive impairment in depression: a systematic review and meta-analysis. Psychol. Med. 44, 2029–2040 (2014).

6. Harvey, P. D. et al. Cognitive dysfunction in schizophrenia: An expert group paper on the current state of the art. Schizophr Res Cogn 29, 100249 (2022).

7. Kirkpatrick, B., Fenton, W. S., Carpenter, W. T., Jr & Marder, S. R. The NIMH-MATRICS consensus statement on negative symptoms. Schizophr. Bull. 32, 214–219 (2006).

8. Correll, C. U. & Schooler, N. R. Negative Symptoms in Schizophrenia: A Review and Clinical Guide for Recognition, Assessment, and Treatment. Neuropsychiatr. Dis. Treat. 16, 519–534 (2020).

9. Lee, J. J. et al. Gene discovery and polygenic prediction from a genome-wide association study of educational attainment in 1.1 million individuals. Nat. Genet. 50, 1112–1121 (2018).

10. Davies, G. et al. Study of 300,486 individuals identifies 148 independent genetic loci influencing general cognitive function. Nat. Commun. 9, 2098 (2018).

11. Lam, M. et al. Collective genomic segments with differential pleiotropic patterns between cognitive dimensions and psychopathology. Nat. Commun. 13, 6868 (2022).

12. Lam, M. et al. Pleiotropic meta-analysis of cognition, education, and schizophrenia differentiates roles of early neurodevelopmental and adult synaptic pathways. Am. J. Hum. Genet. 105, 334–350 (2019).

13. Peyrot, W. J. et al. The association between lower educational attainment and depression owing to shared genetic effects? Results in ~25,000 subjects. Mol. Psychiatry 20, 735–743 (2015).

14. Yuan, S., Xiong, Y., Michaëlsson, M., Michaëlsson, K. & Larsson, S. C. Genetically predicted education attainment in relation to somatic and mental health. Sci. Rep. 11, 4296 (2021).

15. Sivakumaran, S. et al. Abundant pleiotropy in human complex diseases and traits. Am. J. Hum. Genet. 89, 607–618 (2011).

16. Cross-Disorder Group of the Psychiatric Genomics Consortium. Electronic address: & Cross-Disorder Group of the Psychiatric Genomics Consortium. Genomic Relationships, Novel Loci, and Pleiotropic Mechanisms across Eight Psychiatric Disorders. Cell 179, 1469–1482.e11 (2019).

17. Plomin, R. & von Stumm, S. The new genetics of intelligence. Nat. Rev. Genet. 19, 148–159 (2018).

18. Davies, G. et al. Genome-wide association study of cognitive functions and educational attainment in UK Biobank (N=112 151). Mol. Psychiatry 21, 758–767 (2016).

19. Trubetskoy, V. et al. Mapping genomic loci implicates genes and synaptic biology in schizophrenia. Nature 604, 502–508 (2022).

20. Giannakopoulou, O. et al. The Genetic Architecture of Depression in Individuals of East Asian Ancestry: A Genome-Wide Association Study. JAMA Psychiatry 78, 1258–1269 (2021).

21. Chen, T.-T. et al. Shared genetic architectures of educational attainment in East Asian and European populations. Nat Hum Behav 8, 562–575 (2024).

22. Lam, M. et al. Comparative genetic architectures of schizophrenia in East Asian and European populations. Nat. Genet. 51, 1670–1678 (2019).

23. Murphy, A. E., Schilder, B. M. & Skene, N. G. MungeSumstats: a Bioconductor package for the standardization and quality control of many GWAS summary statistics. Bioinformatics 37, 4593–4596 (2021).

24. Turley, P. et al. Multi-trait analysis of genome-wide association summary statistics using MTAG. Nat. Genet. 50, 229–237 (2018).

25. Lee, C. H., Shi, H., Pasaniuc, B., Eskin, E. & Han, B. PLEIO: a method to map and interpret pleiotropic loci with GWAS summary statistics. Am. J. Hum. Genet. 108, 36–48 (2021).

26. Watanabe, K., Taskesen, E., van Bochoven, A. & Posthuma, D. Functional mapping and annotation of genetic associations with FUMA. Nat. Commun. 8, 1826 (2017).

27. 1000 Genomes Project Consortium et al. A global reference for human genetic variation. Nature 526, 68–74 (2015).

28. de Leeuw, C. A., Mooij, J. M., Heskes, T. & Posthuma, D. MAGMA: generalized gene-set analysis of GWAS data. PLoS Comput. Biol. 11, e1004219 (2015).

29. Liberzon, A. et al. The Molecular Signatures Database (MSigDB) hallmark gene set collection. Cell Syst 1, 417–425 (2015).

30. Reimand, J., Kull, M., Peterson, H., Hansen, J. & Vilo, J. g:Profiler--a web-based toolset for functional profiling of gene lists from large-scale experiments. Nucleic Acids Res. 35, W193–200 (2007).

31. Harris, M. A. et al. The Gene Ontology (GO) database and informatics resource. Nucleic Acids Res. 32, D258–61 (2004).

32. Kanehisa, M. & Goto, S. KEGG: kyoto encyclopedia of genes and genomes. Nucleic Acids Res. 28, 27–30 (2000).

33. Bulik-Sullivan, B. et al. An atlas of genetic correlations across human diseases and traits. Nat. Genet. 47, 1236–1241 (2015).

34. Bulik-Sullivan, B. K. et al. LD Score regression distinguishes confounding from polygenicity in genome-wide association studies. Nat. Genet. 47, 291–295 (2015).

35. Zou, Y., Carbonetto, P., Wang, G. & Stephens, M. Fine-mapping from summary data with the ‘Sum of Single Effects’ model. PLoS Genet. 18, e1010299 (2022).

36. Yuan, K. et al. Fine-mapping across diverse ancestries drives the discovery of putative causal variants underlying human complex traits and diseases. Nat. Genet. 56, 1841–1850 (2024).

37. McLaren, W. et al. The Ensembl Variant Effect Predictor. Genome Biol. 17, 122 (2016).

38. Sollis, E. et al. The NHGRI-EBI GWAS Catalog: knowledgebase and deposition resource. Nucleic Acids Res. 51, D977–D985 (2023).

39. Lim, K., Lam, M., Huang, H., Liu, J. & Lee, J. Genetic liability in individuals at ultra-high risk of psychosis: A comparison study of 9 psychiatric traits. PLoS One 15, e0243104 (2020).

40. Euesden, J., Lewis, C. M. & O’Reilly, P. F. PRSice: Polygenic Risk Score software. Bioinformatics 31, 1466–1468 (2015).

41. Choi, S. W. & O’Reilly, P. F. PRSice-2: Polygenic Risk Score software for biobank-scale data. Gigascience 8, (2019).

42. Ge, T., Chen, C.-Y., Ni, Y., Feng, Y.-C. A. & Smoller, J. W. Polygenic prediction via Bayesian regression and continuous shrinkage priors. Nat. Commun. 10, 1776 (2019).

43. Santini, E. & Klann, E. Reciprocal signaling between translational control pathways and synaptic proteins in autism spectrum disorders. Sci. Signal. 7, re10 (2014).

44. Zlomuzica, A., Plank, L. & Dere, E. A new path to mental disorders: Through gap junction channels and hemichannels. Neurosci. Biobehav. Rev. 142, 104877 (2022).

45. Kraft, J. et al. Identifying drug targets for schizophrenia through gene prioritization. Transl. Psychiatry 16, (2026).

46. Tesli, M. et al. VRK2 gene expression in schizophrenia, bipolar disorder and healthy controls. Br. J. Psychiatry 209, 114–120 (2016).

47. Yin, M.-Y. et al. Reduced Vrk2 expression is associated with higher risk of depression in humans and mediates depressive-like behaviors in mice. BMC Med. 21, 256 (2023).

48. Couñago, R. M. et al. Structural characterization of human Vaccinia-Related Kinases (VRK) bound to small-molecule inhibitors identifies different P-loop conformations. Sci. Rep. 7, 7501 (2017).

49. Serafim, R. A. M. et al. Development of pyridine-based inhibitors for the human Vaccinia-related kinases 1 and 2. ACS Med. Chem. Lett. 10, 1266–1271 (2019).

50. Islam, S., Wang, S., Bowden, N., Martin, J. & Head, R. Repurposing existing therapeutics, its importance in oncology drug development: Kinases as a potential target. Br. J. Clin. Pharmacol. 88, 64–74 (2022).

51. Peled, M., Tocheva, A. S., Adam, K. & Mor, A. VRK2 inhibition synergizes with PD-1 blockade to improve T cell responses. Immunol. Lett. 233, 42–47 (2021).

52. Lazo, P. A. VRK2 kinase pathogenic pathways in cancer and neurological diseases. Biochim. Biophys. Acta Mol. Cell Res. 1872, 119949 (2025).

53. Akaneya, Y. et al. Ephrin-A5 and EphA5 interaction induces synaptogenesis during early hippocampal development. PLoS One 5, e12486 (2010).

54. Martínez, A., Otal, R., Sieber, B.-A., Ibáñez, C. & Soriano, E. Disruption of ephrin-A/EphA binding alters synaptogenesis and neural connectivity in the hippocampus. Neuroscience 135, 451–461 (2005).

55. Pasquale, E. B. Eph-ephrin bidirectional signaling in physiology and disease. Cell 133, 38–52 (2008).

56. Pottier, C. et al. Tyrosine kinase inhibitors in cancer: Breakthrough and challenges of targeted therapy. Cancers (Basel) 12, 731 (2020).

57. Orrico, K. B. Basic concepts of cancer genetics and receptor tyrosine kinase inhibition for pharmacists. A narrative review. J. Oncol. Pharm. Pract. 29, 1187–1195 (2023).

58. Staquicini, F. I. et al. First-generation and preclinical evaluation of an EphA5-targeted antibody-drug conjugate in solid tumors. J. Clin. Invest. 135, (2025).

59. Wang, Z. et al. Axon guidance pathway genes are associated with schizophrenia risk. Exp. Ther. Med. 16, 4519–4526 (2018).

60. Notaras, M., Lodhi, A., Fang, H., Greening, D. & Colak, D. The proteomic architecture of schizophrenia iPSC-derived cerebral organoids reveals alterations in GWAS and neuronal development factors. Transl. Psychiatry 11, 541 (2021).

61. Chinnadurai, G., Vijayalingam, S. & Gibson, S. B. BNIP3 subfamily BH3-only proteins: mitochondrial stress sensors in normal and pathological functions. Oncogene 27 Suppl 1, S114– 27 (2008).

62. Kirkin, V. & Rogov, V. V. A diversity of selective autophagy receptors determines the specificity of the autophagy pathway. Mol. Cell 76, 268–285 (2019).

63. Marinković, M. & Novak, I. A brief overview of BNIP3L/NIX receptor-mediated mitophagy. FEBS Open Bio 11, 3230–3236 (2021).

64. Indran, I. R., Tufo, G., Pervaiz, S. & Brenner, C. Recent advances in apoptosis, mitochondria and drug resistance in cancer cells. Biochim. Biophys. Acta 1807, 735–745 (2011).

65. Li, Y. et al. BNIP3L/NIX-mediated mitophagy: molecular mechanisms and implications for human disease. Cell Death Dis. 13, 14 (2021).

66. Gao, A., Jiang, J., Xie, F. & Chen, L. Bnip3 in mitophagy: Novel insights and potential therapeutic target for diseases of secondary mitochondrial dysfunction. Clin. Chim. Acta 506, 72–83 (2020).

67. Li, Y.-Y.Qin, Z.-H. & Sheng, R. The multiple roles of autophagy in neural function and diseases. Neurosci. Bull. 40, 363–382 (2024).

68. Hao, B.-B. et al. The novel cereblon modulator CC-885 inhibits mitophagy via selective degradation of BNIP3L. Acta Pharmacol. Sin. 41, 1246–1254 (2020).

69. Merikangas, A. K. et al. What genes are differentially expressed in individuals with schizophrenia? A systematic review. Mol. Psychiatry 27, 1373–1383 (2022).

70. Zhou, J. et al. Identification of rare and common variants in BNIP3L: a schizophrenia susceptibility gene. Hum. Genomics 14, 16 (2020).

71. Ali, D. et al. Direct targets of MEF2C are enriched for genes associated with schizophrenia and cognitive function and are involved in neuron development and mitochondrial function. PLoS Genet. 20, e1011093 (2024).

72. Dunn, E. C. et al. Genetic determinants of depression: recent findings and future directions. Harv. Rev. Psychiatry 23, 1–18 (2015).

73. Cai, N., Choi, K. W. & Fried, E. I. Reviewing the genetics of heterogeneity in depression: Operationalizations, manifestations, and etiologies. Hum. Mol. Genet. (2020) doi:10.1093/hmg/ddaa115.

74. Liao, S.-C. et al. Low prevalence of major depressive disorder in Taiwanese adults: possible explanations and implications. Psychol. Med. 42, 1227–1237 (2012).

75. Kessler, R. C. & Bromet, E. J. The epidemiology of depression across cultures. Annu. Rev. Public Health 34, 119–138 (2013).

76. Fusar-Poli, P., Yung, A. R., McGorry, P. & van Os, J. Lessons learned from the psychosis high-risk state: towards a general staging model of prodromal intervention. Psychol. Med. 44, 17–24 (2014).

77. McGorry, P. & van Os, J. Redeeming diagnosis in psychiatry: timing versus specificity. Lancet 381, 343–345 (2013).

