## Supplementary Information for "Shared Genetic Architecture of Psychosis, Mood, and Cognition in East Asian Ancestry"

**Quality control of summary statistics**

To ensure consistency of summary statistics for downstream analyses, rigorous quality control (QC) procedures were carried out with the MungeSumstats tool^1^. The default QC parameters within MungeSumstats were applied. These include removing strand ambiguous SNPs and non-biallelic SNPs, ensuring that all variants have non-zero effect sizes and standard errors, and standardizing reference genome build (GRCh37 and dbSNP v.144), SNP ID, and directionality of effect allele. Z-score and effective sample size [Neff = 4 / (1/N_Case_ + 1/N_Control_)], were also calculated with MungeSumstats, for downstream analysis. QC procedures resulted in 5,869,761 SNPs, 6,806,826 SNPs, and 6,087,606 SNPs for the educational attainment, depression and schizophrenia summary statistics respectively (Supplementary Table S1). Heritability estimates were converted from observed to liability scale assuming 6.5% and 1% prevalence for depression and schizophrenia respectively.

**Genome-wide analysis - PLEIO**

For the PLEIO^2^ analysis, the ldsc_preprocess.py script implemented within PLEIO was used to estimate both the genetic covariance and environmental correlation matrices across the three traits. This step also standardises the effect sizes across the summary statistics. To ensure comparability of analytic results with the MTAG analysis, the input SNPs for PLEIO analysis were limited to only include those that were in MTAG analysis. This resulted in 3,183,822 SNPs merged across the three traits for PLEIO analysis. The PLEIO pleiotropic meta-analysis was subsequently implemented with the Pleio.py script. The --blup option (i.e., Best Linear Unbiased Prediction) was also implemented in PLEIO to calculate trait-specific beta values and standard error for each trait. To ensure consistency and comparability with the MTAG results, the GWAS equivalent effective sample size (Neff) for the PLEIO analysis was estimated by utilising the Neff formula as implemented in MTAG [Neff = N_GWAS-Input_ ((χ^2^_PLEIO_-1)/(χ^2^_Input_-1))]^3^. To estimate this, we first converted the P-values from the PLEIO output to Z-score, and from Z-score to mean chi-square. Here, instead of employing a summation of sample size across the three traits, we employed a conservative approach by using the summary statistics with the largest sample size as the input GWAS sample size; in this case it is the summary statistics from educational attainment (N = 176400)^4^.

**Fine-mapping analysis**

To identify 95% credible SNPs from the genome-wide analyses of MTAG and PLEIO, the SuSiEx^5^ approach was conducted. The SuSiEx methodology, which build upon the Sum of Single Effects model (SuSiE^6^), performs fine-mapping for within and cross-population genome-wide significant loci. In situations where SuSiEx was only supplied with one set of input data, it functions as SuSiE; this was implemented in the output for each trait derived from MTAG. For PLEIO output, the trait-specific beta values for educational attainment, depression, and schizophrenia, were entered as a single model in SuSiEx.

**Polygenic risk score analysis**

The predictive ability of the MTAG and PLEIO output were tested in a sample of individuals at risk for psychosis. In the case of PLEIO, as the --blup option only estimates trait-specific beta weights and standard error, a pooled beta weight across the three traits was estimated for the polygenic risk score analysis.

**Pharmacological annotation of statistically prioritized and biologically annotated candidate genes**

The following pharmacological annotation is intended to contextualize candidate genes emerging from the analyses reported in the results and downstream biological annotation. These annotations should be interpreted as exploratory and hypothesis-generating rather than as evidence that the nominated genes are validated therapeutic targets. The primary evidence in the present study is locus-level statistical prioritization from multi-trait genome-wide analyses, fine-mapping, and downstream annotation. Therapeutic interpretation requires additional experimental validation, including causal variant identification, tissue- and isoform-specific functional studies, target engagement assays, and safety assessment.

**VRK2**

*VRK2* encodes a serine/threonine kinase and is therefore part of a target class with substantial precedent for pharmacological modulation. Prior gene-prioritization work has nominated VRK2 as a potentially druggable target in schizophrenia^7^, and the present findings extend its relevance to the broader affective–psychosis genetic architecture examined in East Asian ancestry. From a pharmacological perspective, *VRK2* is structurally attractive because it belongs to the kinase family, where ATP-binding pockets, hinge-region interactions, and activation-loop dynamics have historically supported medicinal chemistry development^8–10^. Existing structural work on vaccinia-related kinases, including inhibitor-bound conformations and VRK-family P-loop dynamics, provides a basis for future probe development and structure-guided optimization^8,9^.

However, caution is warranted in interpreting *VRK2* as therapeutic. The statistical-genetics evidence nominates *VRK2* as a candidate locus but, by itself, does not establish whether increasing, decreasing, or otherwise modulating *VRK2* activity would be clinically beneficial. Expression-based evidence suggests that altered or reduced *VRK2/Vrk2* expression may be relevant to schizophrenia, bipolar disorder, and depression-related phenotypes^11,12^. This may point toward a reduced-expression or loss-of-function hypothesis in some contexts, which is not automatically aligned with conventional kinase-inhibitor strategies. Additional evidence from cancer and immune biology supports the broader biological relevance of *VRK2* and demonstrates that *VRK2* modulation can have functional consequences in other disease contexts^13,14^. Therefore, *VRK2* should be presented as a pharmacologically tractable candidate for functional follow-up, not as an immediately actionable inhibitory target.

**EPHA5**

*EPHA5* encodes an *EphA5* receptor tyrosine kinase involved in Eph/ephrin signaling. This pathway has established roles in axon guidance, neuronal migration, synaptogenesis, synaptic plasticity, neural connectivity, and circuit formation during neurodevelopment^15–17^. These biological functions are relevant to psychiatric genetic findings because neurodevelopmental and synaptic organization pathways are recurrently implicated in schizophrenia and related phenotypes^18,19^. *EPHA5*, therefore, provides a plausible biological bridge between statistical genetic prioritization and neurodevelopmental mechanisms.

Pharmacologically, *EPHA5* is notable because receptor tyrosine kinases are a well-developed therapeutic class in oncology^20,21^. The extracellular ligand-binding domain may be accessible to biologics, while the intracellular kinase domain provides a potential small-molecule target. Oncology programs targeting Eph-family receptors, including antibody-drug conjugate approaches, illustrate the tractability of this target class in non-psychiatric disease contexts^22^. Nonetheless, this oncology precedent should not be overinterpreted for psychiatry. CNS translation faces additional constraints, including blood-brain barrier penetration, timing during neurodevelopment, cell-type specificity, and the risk of perturbing broadly important guidance pathways. At present, *EPHA5* is best framed as a biologically interpretable neurodevelopmental candidate with pharmacological precedent in other disease areas, rather than as a near-term psychiatric drug target.

**BNIP3L**

*BNIP3L*, also known as *NIX*, has a different pharmacological profile from *VRK2* and *EPHA5*. It encodes a mitochondrial outer-membrane protein involved in mitophagy, mitochondrial quality control, mitochondrial stress responses, and stress-induced apoptosis^23–26^. These processes are relevant to psychiatric biology insofar as mitochondrial dysfunction, oxidative stress, altered proteostasis, and autophagy-related mechanisms have been implicated in neural function and neuropsychiatric disease biology^26–31^. BNIP3L, therefore, provides a potential link between genetic findings and mitochondrial pathway biology.

Unlike kinases or receptor tyrosine kinases, *BNIP3L* does not present an obvious enzymatically active site or canonical kinase-like ATP-binding pocket. Direct small-molecule targeting is therefore likely to be more challenging. Its pharmacological relevance is more plausibly pathway-level: modulation of mitophagy, proteostasis, mitochondrial stress responses, or targeted degradation mechanisms may provide indirect routes for experimental perturbation^26–28,32^. For example, *CC-885* has been reported to inhibit mitophagy by selectively degrading *BNIP3L**^32^*, demonstrating one potential experimental approach to perturb this pathway. Genetic and transcriptomic evidence also supports a possible relationship between *BNIP3L* and schizophrenia-relevant biology, including reports of rare and common *BNIP3L* variants associated with schizophrenia susceptibility and broader evidence of altered gene expression in schizophrenia^29,30^. In addition, *MEF2C* target-gene analyses implicate neurodevelopmental and mitochondrial pathways relevant to schizophrenia and cognitive function, suggesting a broader mechanism at play^31^.

The directionality of *BNIP3L* modulation remains uncertain. Mitophagy can be protective or maladaptive depending on cellular context, disease stage, stress exposure, and tissue compartment^26–28^. Therefore, any proposal to inhibit, activate, or degrade *BNIP3L* requires direct functional validation in disease-relevant neuronal and glial systems. *BNIP3L* is best interpreted as a candidate for pathway-level experimental follow-up rather than as a conventional, directly druggable psychiatric target.

**Summary**

Together, these annotations suggest a hierarchy of pharmacological interpretability among the nominated genes. *VRK2* is the most conventionally tractable because it is a kinase with existing structural and chemical-biology precedent. *EPHA5* is biologically plausible and belongs to a drugged receptor tyrosine kinase class, but psychiatric translation remains speculative and would require CNS-specific validation. *BNIP3L* highlights mitochondrial and mitophagy biology, with potential for pathway-level modulation rather than straightforward direct targeting. These observations support future functional and pharmacological studies but should not be interpreted as evidence of therapeutic readiness.

**Data and code availability**

1. Publicly available GWAS summary statistics
   a. Educational attainment: <https://www.ebi.ac.uk/gwas/studies/GCST90296498>
   b. Depression: <https://pgc.unc.edu/for-researchers/download-results/>
   c. Schizophrenia: <https://pgc.unc.edu/for-researchers/download-results/>
2. Bioinformatics tool
   a. MungeSumstats: <https://github.com/neurogenomics/MungeSumstats>
   b. MTAG: <https://github.com/JonJala/mtag>
   c. PLEIO: <https://github.com/cuelee/pleio>
   d. FUMA: <https://fuma.ctglab.nl/>
   e. LDSC: <https://github.com/bulik/ldsc>
   f. SuSiEx: <https://github.com/getian107/SuSiEx>
   g. VEP: <https://grch37.ensembl.org/Homo_sapiens/Tools/VEP>
   h. GWAS catalog: <https://www.ebi.ac.uk/gwas/search>
   i. g:Profiler: <https://biit.cs.ut.ee/gprofiler/gost>
   j. PRSice2: <https://choishingwan.github.io/PRSice/>
   k. PRS-CS: <https://github.com/getian107/PRScs>

**References**

1.	Murphy, A. E., Schilder, B. M. & Skene, N. G. MungeSumstats: a Bioconductor package for the standardization and quality control of many GWAS summary statistics. *Bioinformatics* **37**, 4593–4596 (2021).

2.	Lee, C. H., Shi, H., Pasaniuc, B., Eskin, E. & Han, B. PLEIO: a method to map and interpret pleiotropic loci with GWAS summary statistics. *Am. J. Hum. Genet.* **108**, 36–48 (2021).

3.	Turley, P. *et al.* Multi-trait analysis of genome-wide association summary statistics using MTAG. *Nat. Genet.* **50**, 229–237 (2018).

4.	Chen, T.-T. *et al.* Shared genetic architectures of educational attainment in East Asian and European populations. *Nat Hum Behav* **8**, 562–575 (2024).

5.	Yuan, K. *et al.* Fine-mapping across diverse ancestries drives the discovery of putative causal variants underlying human complex traits and diseases. *medRxiv* (2023) doi:[10.1101/2023.01.07.23284293](http://dx.doi.org/10.1101/2023.01.07.23284293).

6.	Wang, G., Sarkar, A., Carbonetto, P. & Stephens, M. A simple new approach to variable selection in regression, with application to genetic fine mapping. *J. R. Stat. Soc. Series B Stat. Methodol.* **82**, 1273–1300 (2020).

7.	Kraft, J. *et al.* Identifying drug targets for schizophrenia through gene prioritization. *Transl. Psychiatry* **16**, (2026).

8.	Couñago, R. M. *et al.* Structural characterization of human Vaccinia-Related Kinases (VRK) bound to small-molecule inhibitors identifies different P-loop conformations. *Sci. Rep.* **7**, 7501 (2017).

9.	Serafim, R. A. M. *et al.* Development of pyridine-based inhibitors for the human Vaccinia-related kinases 1 and 2. *ACS Med. Chem. Lett.* **10**, 1266–1271 (2019).

10.	Islam, S., Wang, S., Bowden, N., Martin, J. & Head, R. Repurposing existing therapeutics, its importance in oncology drug development: Kinases as a potential target. *Br. J. Clin. Pharmacol.* **88**, 64–74 (2022).

11.	Tesli, M. *et al.* VRK2 gene expression in schizophrenia, bipolar disorder and healthy controls. *Br. J. Psychiatry* **209**, 114–120 (2016).

12.	Yin, M.-Y. *et al.* Reduced Vrk2 expression is associated with higher risk of depression in humans and mediates depressive-like behaviors in mice. *BMC Med.* **21**, 256 (2023).

13.	Peled, M., Tocheva, A. S., Adam, K. & Mor, A. VRK2 inhibition synergizes with PD-1 blockade to improve T cell responses. *Immunol. Lett.* **233**, 42–47 (2021).

14.	Lazo, P. A. VRK2 kinase pathogenic pathways in cancer and neurological diseases. *Biochim. Biophys. Acta Mol. Cell Res.* **1872**, 119949 (2025).

15.	Akaneya, Y. *et al.* Ephrin-A5 and EphA5 interaction induces synaptogenesis during early hippocampal development. *PLoS One* **5**, e12486 (2010).

16.	Martínez, A., Otal, R., Sieber, B.-A., Ibáñez, C. & Soriano, E. Disruption of ephrin-A/EphA binding alters synaptogenesis and neural connectivity in the hippocampus. *Neuroscience* **135**, 451–461 (2005).

17.	Pasquale, E. B. Eph-ephrin bidirectional signaling in physiology and disease. *Cell* **133**, 38–52 (2008).

18.	Wang, Z. *et al.* Axon guidance pathway genes are associated with schizophrenia risk. *Exp. Ther. Med.* **16**, 4519–4526 (2018).

19.	Notaras, M., Lodhi, A., Fang, H., Greening, D. & Colak, D. The proteomic architecture of schizophrenia iPSC-derived cerebral organoids reveals alterations in GWAS and neuronal development factors. *Transl. Psychiatry* **11**, 541 (2021).

20.	Pottier, C. *et al.* Tyrosine kinase inhibitors in cancer: Breakthrough and challenges of targeted therapy. *Cancers (Basel)* **12**, 731 (2020).

21.	Orrico, K. B. Basic concepts of cancer genetics and receptor tyrosine kinase inhibition for pharmacists. A narrative review. *J. Oncol. Pharm. Pract.* **29**, 1187–1195 (2023).

22.	Staquicini, F. I. *et al.* First-generation and preclinical evaluation of an EphA5-targeted antibody-drug conjugate in solid tumors. *J. Clin. Invest.* **135**, (2025).

23.	Chinnadurai, G., Vijayalingam, S. & Gibson, S. B. BNIP3 subfamily BH3-only proteins: mitochondrial stress sensors in normal and pathological functions. *Oncogene* **27 Suppl 1**, S114–27 (2008).

24.	Kirkin, V. & Rogov, V. V. A diversity of selective autophagy receptors determines the specificity of the autophagy pathway. *Mol. Cell* **76**, 268–285 (2019).

25.	Marinković, M. & Novak, I. A brief overview of BNIP3L/NIX receptor-mediated mitophagy. *FEBS Open Bio* **11**, 3230–3236 (2021).

26.	Li, Y. *et al.* BNIP3L/NIX-mediated mitophagy: molecular mechanisms and implications for human disease. *Cell Death Dis.* **13**, 14 (2021).

27.	Gao, A., Jiang, J., Xie, F. & Chen, L. Bnip3 in mitophagy: Novel insights and potential therapeutic target for diseases of secondary mitochondrial dysfunction. *Clin. Chim. Acta* **506**, 72–83 (2020).

28.	Li, Y.-Y., Qin, Z.-H. & Sheng, R. The multiple roles of autophagy in neural function and diseases. *Neurosci. Bull.* **40**, 363–382 (2024).

29.	Merikangas, A. K. *et al.* What genes are differentially expressed in individuals with schizophrenia? A systematic review. *Mol. Psychiatry* **27**, 1373–1383 (2022).

30.	Zhou, J. *et al.* Identification of rare and common variants in BNIP3L: a schizophrenia susceptibility gene. *Hum. Genomics* **14**, 16 (2020).

31.	Ali, D. *et al.* Direct targets of MEF2C are enriched for genes associated with schizophrenia and cognitive function and are involved in neuron development and mitochondrial function. *PLoS Genet.* **20**, e1011093 (2024).

32.	Hao, B.-B. *et al.* The novel cereblon modulator CC-885 inhibits mitophagy via selective degradation of BNIP3L. *Acta Pharmacol. Sin.* **41**, 1246–1254 (2020).
