## Supplementary Figures for "Shared Genetic Architecture of Psychosis, Mood, and Cognition in East Asian Ancestry"

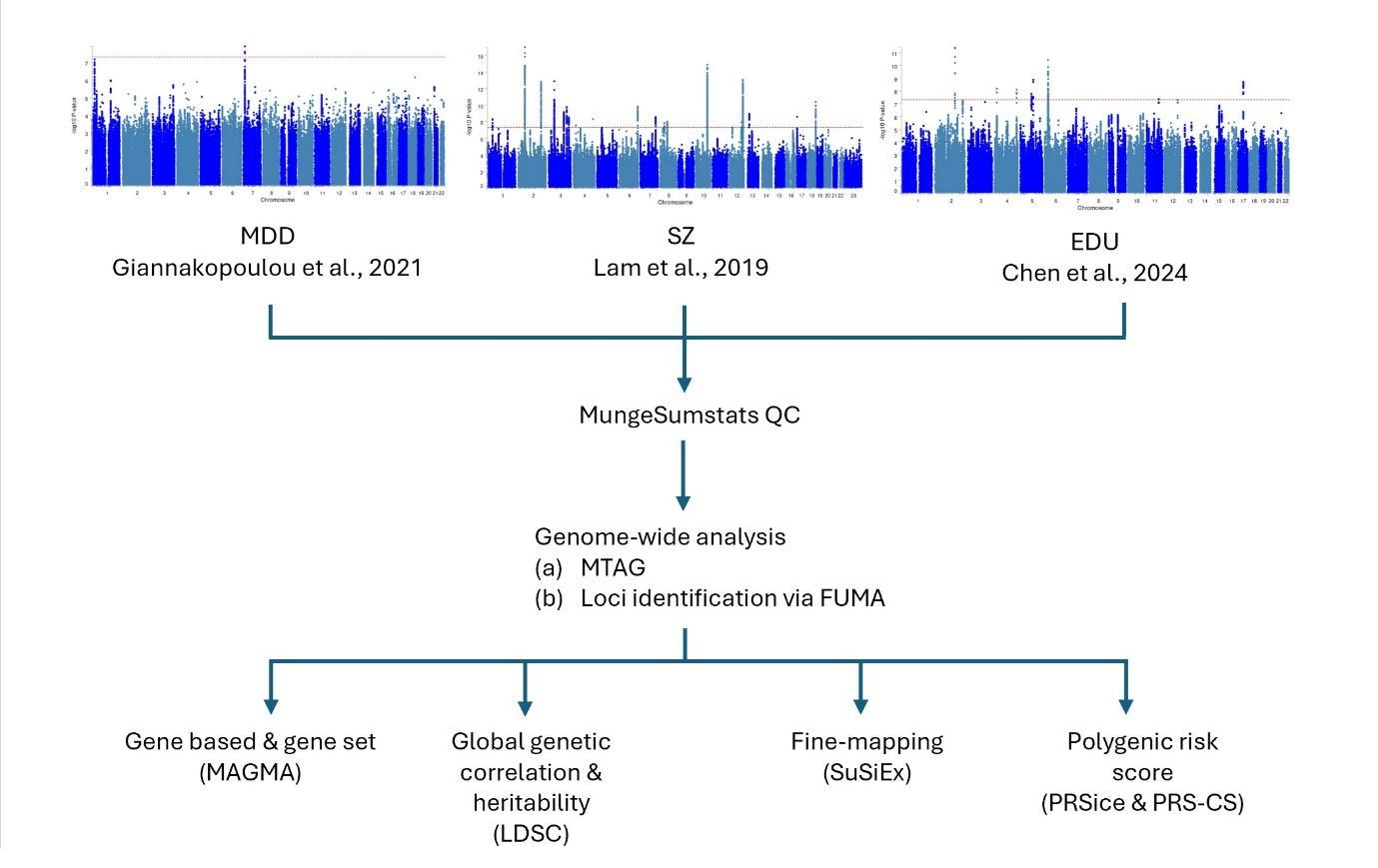

**Supplementary Figure 1. Flow chart of analyses.**

*Note*. MDD = Depression; SZ = Schizophrenia; EDU = Educational attainment.

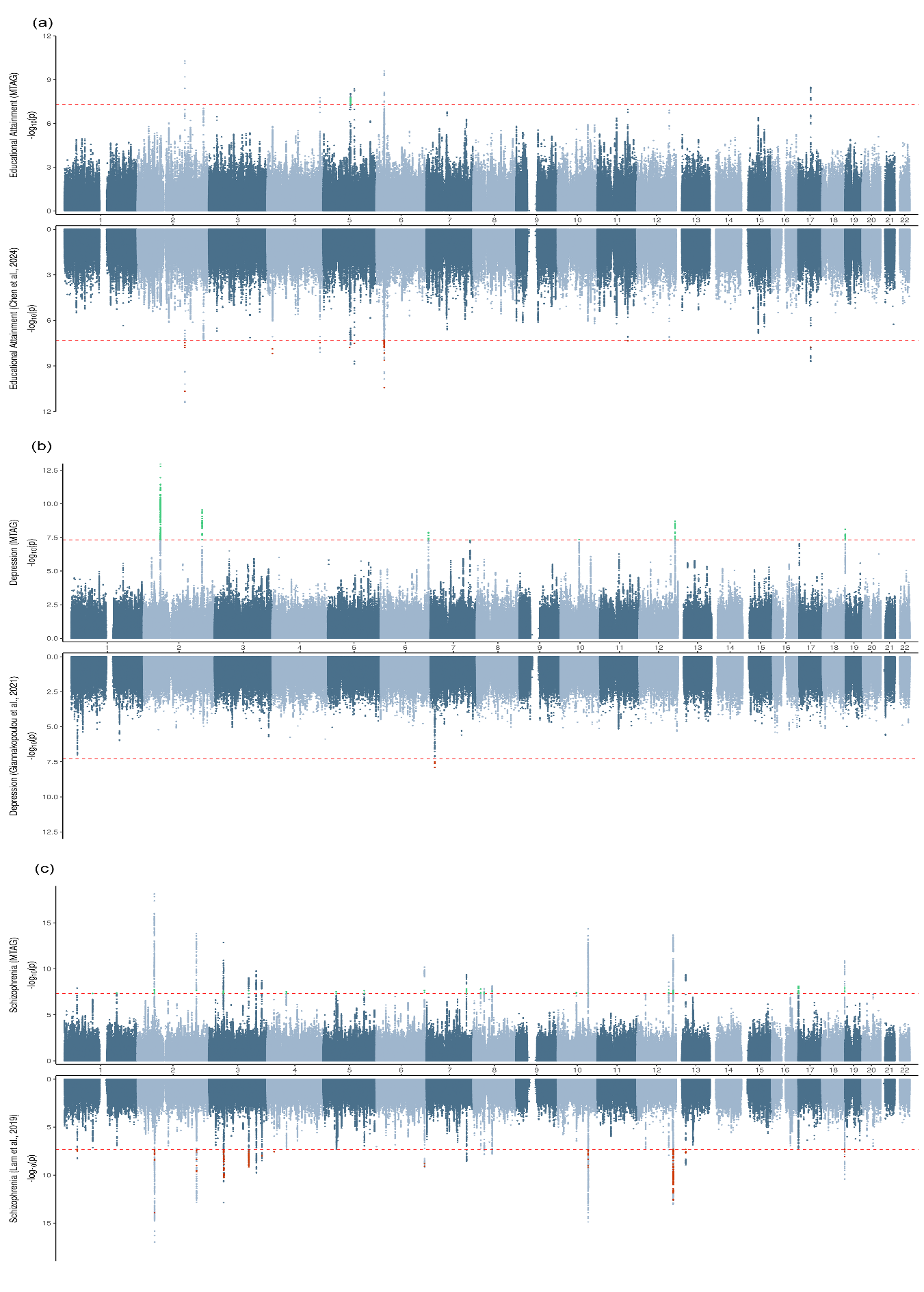

**Supplementary Figure 2. Miami plot of educational attainment, depression and schizophrenia**

*Note*. Top panel: Visualisation of genome-wide association for MTAG output for (a) Educational attainment; (b) Depression; (c) Schizophrenia. Bottom panel: Visualisation of genome-wide association for input dataset by (a) Educational attainment - Chen et al., (2024); (b) Depression - Giannakopoulou et al., (2021); (c) Schizophrenia - Lam et al., (2019). Novel loci are represented by green coloured dots. Loci that did not reached genome-wide significance, compared to the input dataset, are represented by orange coloured dots. The red dotted line represents genome-wide significance of p = 5x10^-8^.

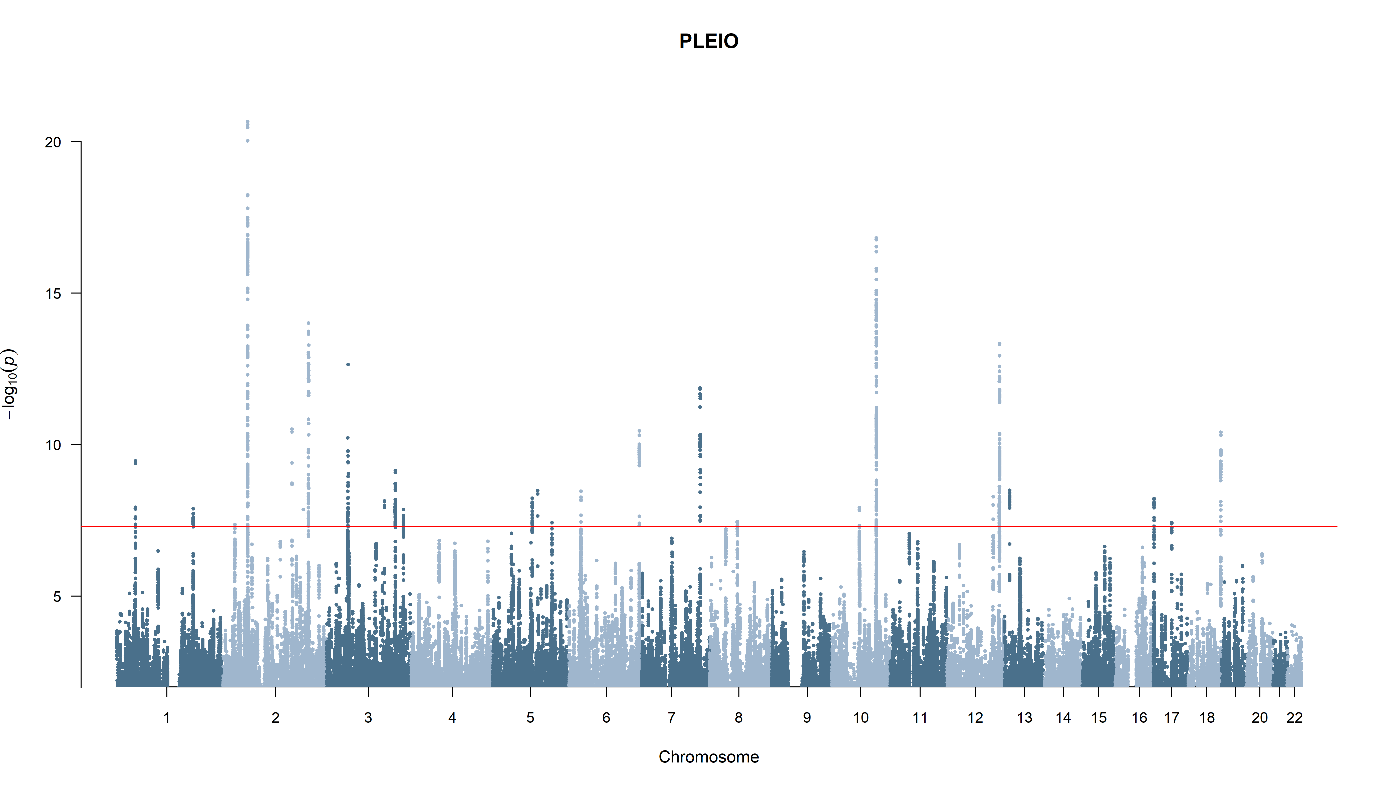

**Supplementary Figure 3. Manhattan plot of PLEIO analysis**

*Note.* The red line represents genome-wide significance of p = 5x10^-8^.

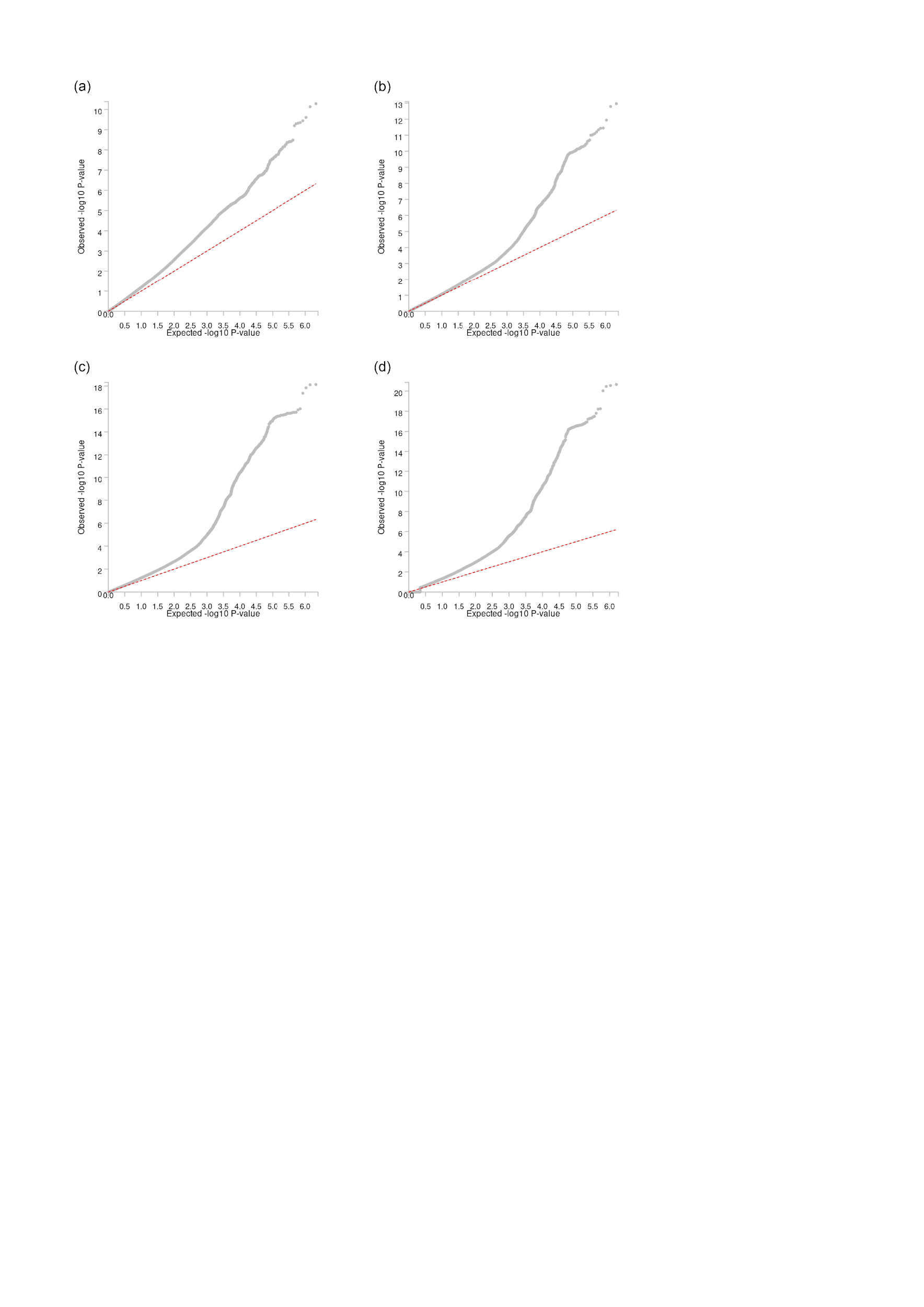

**Supplementary Figure 4. QQ Plots for MTAG and PLEIO analysis**

Panel (a) QQ plot for MTAG educational attainment; (b) QQ plot for MTAG depression; (c) QQ plot for MTAG schizophrenia; (d) QQ plot for PLEIO.

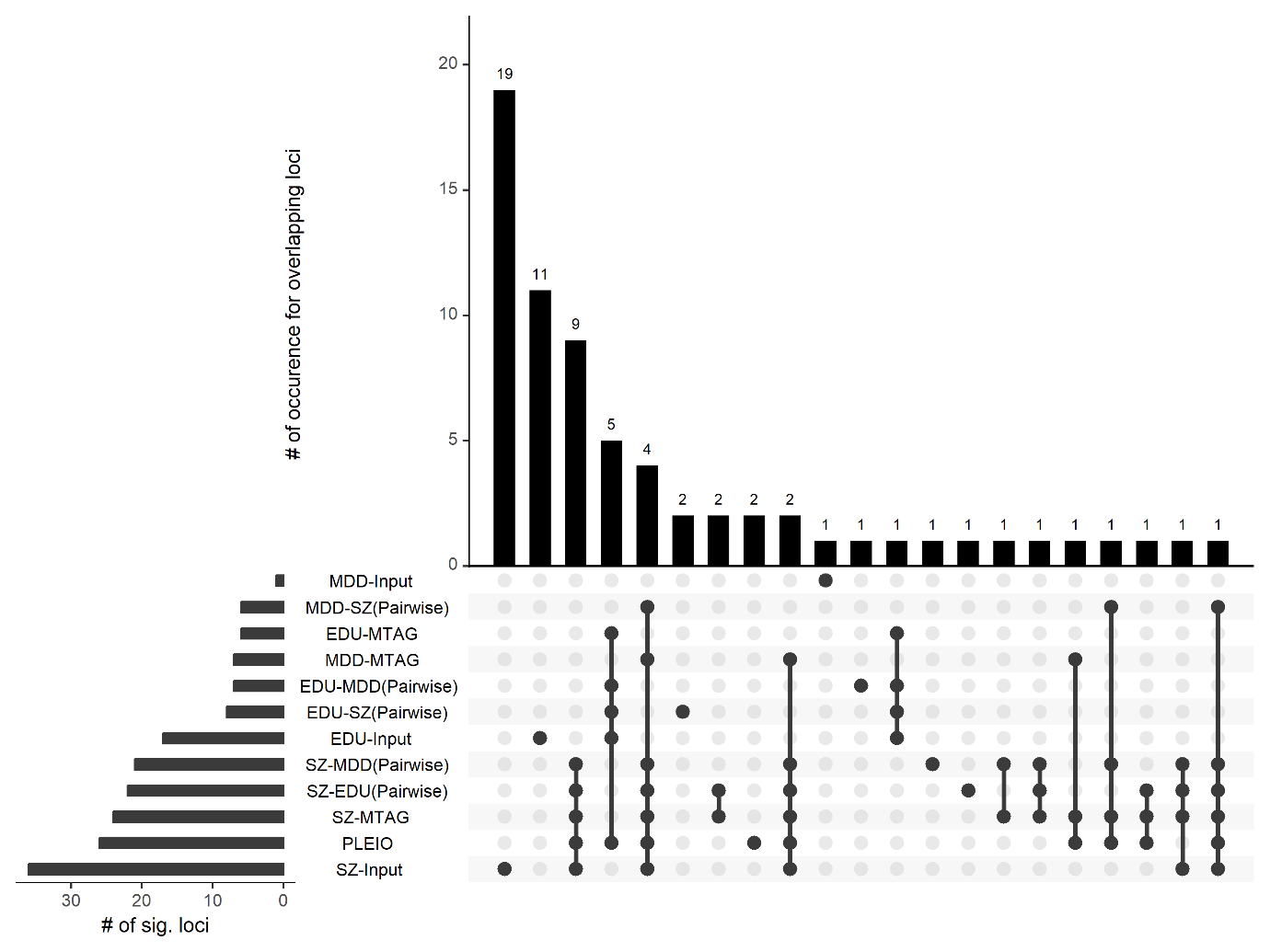

**Supplementary Figure 5. Visualisation for overlapping genome-wide significant loci between each trait – Pairwise and 3-trait MTAG model**

*Note.* The traits listed refers to 3-way MTAG model, unless otherwise stated. EDU-MTAG = Educational attainment MTAG output; MDD-MTAG = Depression MTAG output; SZ-MTAG = Schizophrenia MTAG output; EDU-Input = Discovery input dataset by Chen et al. (2024); SZ-Input = Discovery input dataset by Lam et al. (2019).

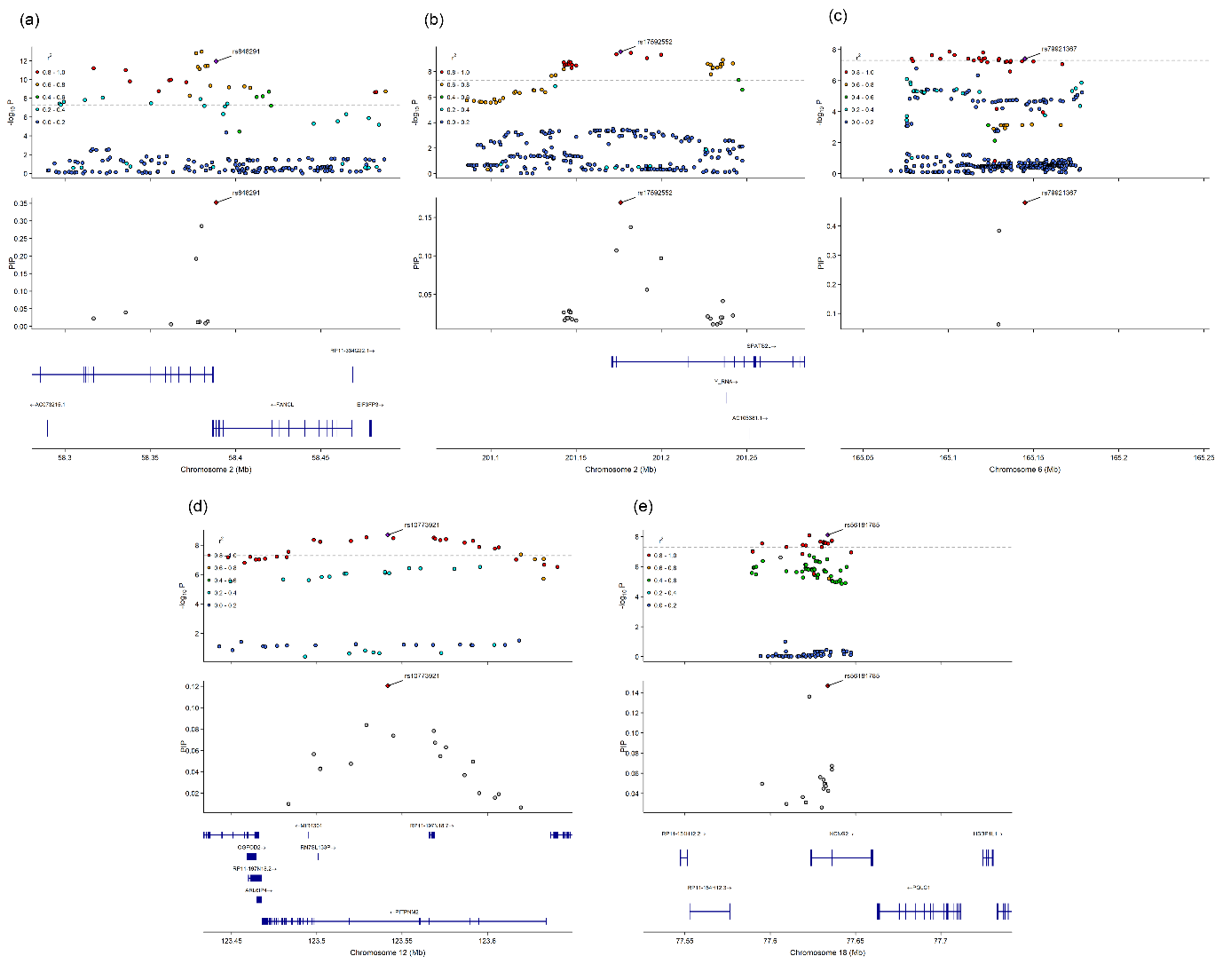

**Supplementary Figure 6. Regional plot and fine-mapping posterior inclusion probability for novel locus detected in MTAG Depression**

The top panel is a regional plot made with LocuszoomR; the locus start and end position is as defined by the MTAG-GWAS results. The middle panel shows posterior inclusion probability results from SuSiEx. Panel (a) MDD-MTAG locus 5; (b) MDD-MTAG locus 8; (c) MDD-MTAG locus 20; (d) MDD-MTAG locus 28; (e) MDD-MTAG locus 32.

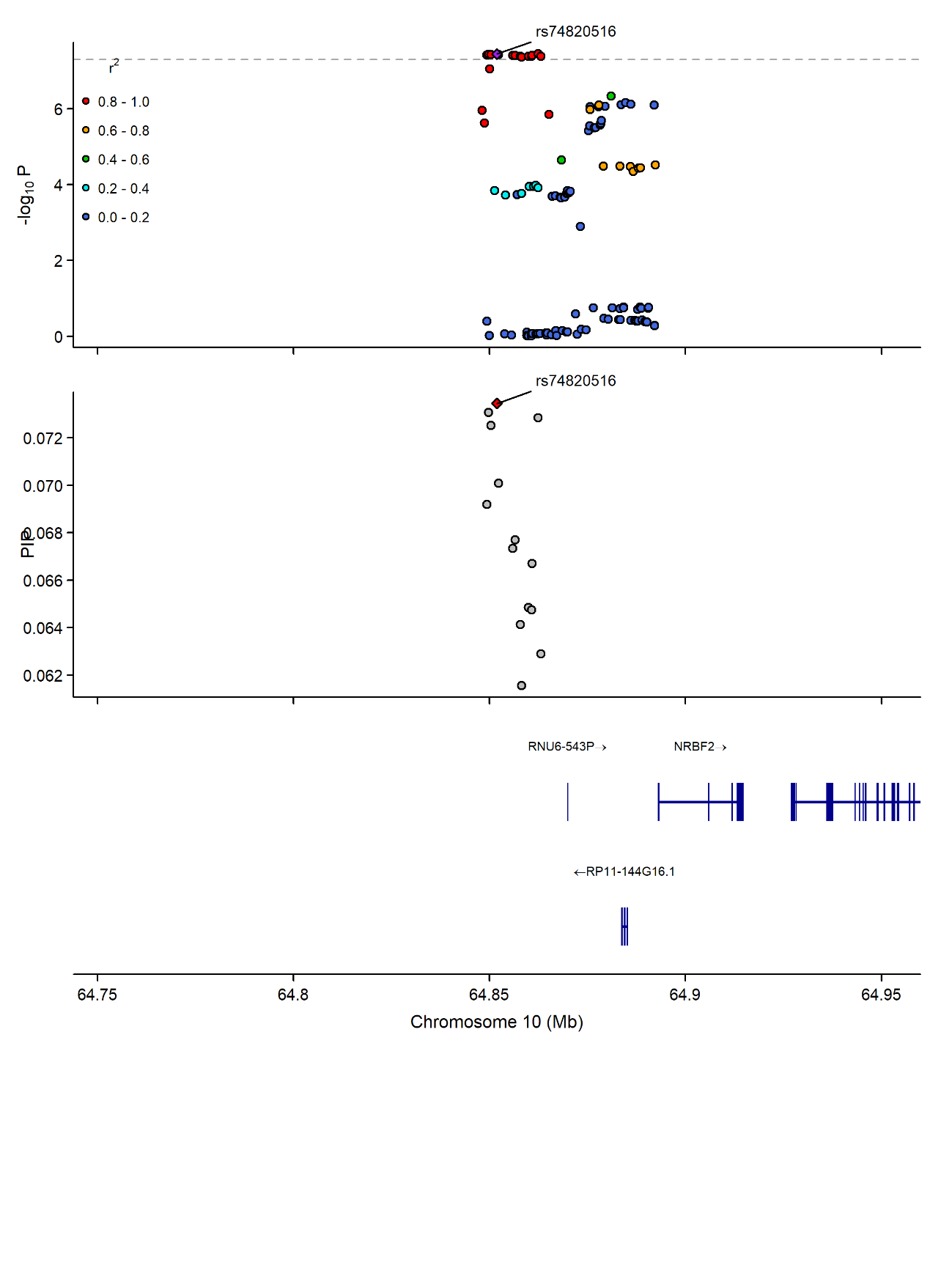

**Supplementary Figure 7. Regional plot and fine-mapping posterior inclusion probability for novel locus detected in MTAG Schizophrenia**

The top panel is a regional plot made with LocuszoomR; the locus start and end position is as defined by the MTAG-GWAS results. The middle panel shows posterior inclusion probability results from SuSiEx. The result presented is for SZ-MTAG locus 25.

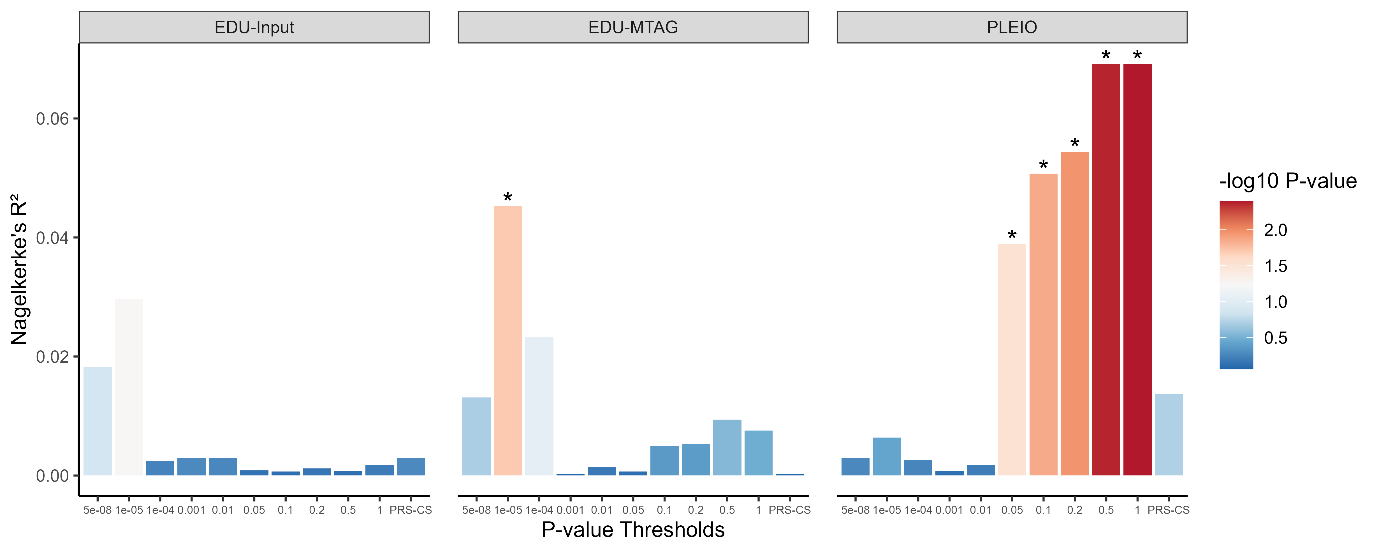

**Supplementary Figure 8. Polygenic prediction in individuals at ultra risk for psychosis versus healthy controls**

*Note.* EDU-Input = The discovery input dataset by Chen et al., (2024), with quality control procedures applied via MungeSumstats; EDU-MTAG = Educational attainment MTAG output. *p < 0.05.
